# Toxicity-specific peripheral blood T and B cell dynamics in anti-PD-1 and combined immune checkpoint inhibition

**DOI:** 10.1101/2023.01.20.23284818

**Authors:** Mick J.M. van Eijs, Rik J. Verheijden, Stefanie A. van der Wees, Stefan Nierkens, Anne S.R. van Lindert, Karijn P.M. Suijkerbuijk, Femke van Wijk, the UNICIT Consortium

## Abstract

Immune checkpoint inhibitors (ICI) have revolutionized the treatment landscape of advanced malignancies, but come with a diverse spectrum of immune-related adverse events (irAEs). Mechanistic studies can aid the transition from expert-opinion to evidence-based irAE treatment strategies. We aimed to longitudinally characterize peripheral blood T and B cell dynamics in ICI-treated patients by multicolor flow cytometry and serum multiplex immunoassay at baseline, ±3 weeks and ±6 weeks or upon clinically relevant irAEs. We analyzed samples from 44 ICI-treated patients (24 anti-PD-1 monotherapy, 20 combined anti-PD-1/anti-CTLA-4; cICI), of whom 21 developed irAEs, and 10 healthy donors. IrAEs after cICI were characterized by significantly enhanced proliferation of Th1-associated, mainly (CD4^+^) effector memory T cells, as well as Th17-associated immune responses and germinal center activation (reflected by CXCL13 and IL-21 increases). We observed no changes in CD21^lo^, memory, class-switched or newly activated B cell subsets. Especially double-positive PD-1^+^LAG-3^+^ CD8^+^ T cells showed enhanced cytotoxic capacity in patients with irAEs after cICI. Within anti-PD-1 monotherapy, irAEs were associated with modestly enhanced Th1-associated responses reflected by increased serum CXCL9 and CXCL10. In conclusion, ICI-induced toxicity is dominated by enhanced Th1-associated responses, but in cICI we also found evidence for Th17-associated responses and germinal center activation. Together, our data add to the growing body of evidence that irAEs may be driven by newly activated CD4^+^ helper T cells, specifically after cICI. This study also supports tailored irAE treatment, based on ICI regimen, and to deploy specific strategies such as Th17 inhibition especially in cICI-associated irAEs.

## Introduction

Immune checkpoint inhibitors (ICI) have realized unprecedented survival improvements in a selection of patients with advanced malignancies, but comes with a broad spectrum of immune-related adverse events (irAEs) [1]. To date, recommendations for irAE management are largely based on multidisciplinary expert consensus [2-4]. In quest of rationally developed treatment strategies, it has been suggested to draw on experience with specific autoimmune diseases (AD) these irAEs mimic, assuming shared pathophysiology [1]. Despite similarities, immune-related toxicity should clinically be considered a separate disease entity, for instance illustrated by the more acute onset and possibility of complete irAE reversibility with adequate therapy. Together with the concern that targeted immunosuppression may thwart antitumor immunity [5, 6], these observations underscore that evidence-based irAE treatment requires more profound biological understanding of specific mechanisms underlying ICI-induced toxicity.

Many efforts have been undertaken to predict irAEs at baseline or early on-treatment based on peripheral blood immune cells, cytokines, auto-antibodies and gut microbiome, as extensively reviewed in Hommes et al. [7]. Relatively higher blood lymphocyte count at baseline has been associated with development of irAEs [7]. Several studies that further characterized T and B cells at baseline and on-treatment reported a multitude of factors associated with irAE development, such as proliferation of (activated) CD4^+^ and CD8^+^ T cell subsets, baseline Th17 dominance and early increase in CD21^lo^ B cells [8-12]. Most of these findings still warrant replication. Particularly effector memory CD4^+^ T cells (CD4_EM_) have recently been suggested as drivers of irAEs [10, 13, 14]. CD4^+^ T cell clonal expansion preceding irAEs [10, 15] and indications for epitope sharing between tumor and irAE-affected tissue [16, 17] suggests a role for T cell auto-reactivity in irAE development [10, 13]. However, the concept of genuine autoimmunity does not seem to completely recapitulate the pathophysiology of irAEs.

Longitudinal studies into general irAE mechanisms comparing different ICI regimens are lacking, but urgently needed to improve irAE treatment strategies [18]. Besides, the role of B cells in irAEs has received little attention. Therefore, we interrogated peripheral T and B cell dynamics and cytokine production from baseline to several weeks on-treatment and upon immune-related toxicity in anti-PD-1 monotherapy and combined anti-CTLA-4 and anti-PD-1 (cICI) treated patients.

## Methods

### Study population and design

We included patients from the UNICIT biobank study conducted within the University Medical Center Utrecht (UMCU), Utrecht, the Netherlands. Adult patients undergoing a first regimen of ICI with anti-PD-1 monotherapy, combined anti-CTLA-4 and anti-PD-1 (cICI), or anti-PD-(L)1 in combination with chemotherapy for a solid malignancy are prospectively enrolled in this biobank. Stool and blood samples are collected at baseline, 3-4 weeks and 6-8 weeks after treatment initiation, and upon onset of irAEs prior to initiation of immunosuppressive therapy. Furthermore, stool and blood samples are collected after every new line immunosuppression administered for irAEs. If patients undergo diagnostic procedures for suspected immune-mediated toxicity, extra biopsies of irAE affected tissue are obtained. Tumor response to ICI per RECIST 1.1 [19], irAE incidence and severity per CTCAE version 5 [20], as well as doses and duration of all immunosuppressives administered for irAEs are continuously recorded.

Samples from patients treated with cICI or anti-PD-1 monotherapy (without chemotherapy) were used in the present study. We included peripheral blood mononuclear cell (PBMC) and serum samples collected at baseline, at 3-4 weeks, and at 6-8 weeks for patients without (NOTx) or at irAE onset for patients with clinically relevant toxicity (TOX). ‘Clinically relevant toxicity’ was defined as CTCAE v5 grade ≥2 irAEs leading to 1) temporary or permanent ICI discontinuation, and 2) demanding hospitalization and/or ≥0.5 mg/kg daily prednisone as first-line immunosuppression. Healthy donor PBMCs were obtained from the UMCU Mini Donor Service (MDS).

### Blood sample processing and flow cytometry

PBMCs were isolated using Ficoll-Paque density-gradient centrifugation (GE Healthcare), frozen in RPMI-1640 medium (Gibco) supplemented with 2 mM L-glutamine (L-glu; Gibco), 100 IU/ml penicillin-streptomycin (p/s; Gibco), 20% fetal bovine serum (FBS; Invitrogen) and 10% DMSO (Sigma-Aldrich) and stored at -180°C. Serum was isolated and frozen at -80°C within 4 hours after blood collection.

Thawed PBMCs were plated at 0.5–1.0×10^6^ cells per well. Prior to staining, cytokine-containing panels were restimulated with 20 ng/ml phorbol 12-myristate 13-actetate (PMA; Sigma-Aldrich) and 1.0 μg/ml ionomycin (Sigma-Aldrich; 4h, 37°C, 5% CO_2_). Golgistop (0.26% monensin; BD Biosciences, 1:1,500) was added for the last 3.5h. Cells were first incubated (30min., 4°C) with eBioscience Fixable Viability Dye eFluor506 (Invitrogen), washed and then incubated with the surface antibody mix (**Supplementary Table 1**) in FACS buffer (phosphate buffered saline [PBS, Sigma-Aldrich], 2% FBS, 0.1% sodium azide; 25min., 4°C). On-treatment PD-1 expression was measured indirectly using biotinylated anti-human IgG4 alone (for unstimulated) or combined with anti-PD-1 (for stimulated panels) [21], followed by secondary staining with BV711-conjugated streptavidin (30min., 4°C). After fixation and permeabilization with eBioscience fixation/permeabilization reagent (Invitrogen; 30min., 4°C) and washing, cells were incubated (30min., 4°C) with the intracellular/intranuclear antibody mix (**Supplementary Table 1**) and then measured the same or next day on an LSR Fortessa. FlowJo software was used to analyze data; gating strategies are in **Supplementary Fig. 5a-b**.

### Multiplex immunoassay

Concentrations of 23 soluble factors (**Supplementary Table 2**) were measured in serum collected at the same timepoints as PBMCs by an in-house developed and validated immunoassay based on Luminex technology (xMAP, Luminex) at the Multiplex Core Facility (UMCU) [22, 23]. Aspecific heterophilic immunoglobulins were preabsorbed with heteroblock (Omega Biologicals) and acquisition was performed with the Biorad FlexMAP3D (Biorad laboratories) in combination with xPONENT software (v4.2, Luminex). Data was analyzed by 5-parametric curve fitting using Bio-Plex Manager software (v6.1.1, Biorad). Serum samples from 10 healthy donors other than PBMC healthy donors were obtained from the UMCU MDS (median age 35 years, range 26-61; 60% male sex). For one patient no serum was available. Undetectable values below the limit-of-detection (LOD) were replaced with 0.5×LOD.

### Unsupervised and statistical analysis

Analyses were performed with R version 4.2.0. For principal component analysis (PCA), missing data were imputed with the population median per parameter and PCA was performed using *prcomp()* in *stats* (v3.6.2) on all 159 read-out parameters across all panels. t-Distributed stochastic neighbor embedding (t-SNE) projections for panel 2 (**Supplementary Table 1**) were created using *Rtsne* (θ=0.5; v0.16) and k-means clustered self-organizing maps (SOMs) using FlowSOM (v2.4.0) [24] with a random 10% subset of pre-gated alive CD3^+^ singlets drawn from samples upon timepoint 3, toxicity or healthy donor. Longitudinal continuous data were analyzed by linear mixed effects models using the *nlme* package (v3.1-158). Random intercept for subject models with fixed effects for ICI regimen, toxicity status and their interaction with time were fit by restricted maximum likelihood. Significance for fixed effects was evaluated with Satterthwaite approximations given relatively small sample size [25]. Model output was visualized as predicted population estimates with 95% confidence intervals (CIs). Mean and 95% CI (by one-sample t-test) for healthy donor values was plotted as solid and dashed grey lines, respectively. Continuous variables were analyzed with one-sample t tests for single groups, or a Wilcoxon rank sum test (for unpaired) or Wilcoxon signed-rank test (for paired data) between two groups. More than two groups were compared with the Kruskal-Wallis test followed by Dunn’s post-hoc test with Benjamini-Hochberg false discovery rate correction. *P* values <0.05 (two-sided) were considered statistically significant.

### Ethics approval

The UNICIT biobank study was not considered subject to the Dutch Medical Research with Human Subjects Law by the medical research ethics committee of the UMCU. The biobank review committee of the UMCU approved the UNICIT biobank protocol (TCbio 18-123) and granted permission for use of human biospecimens for the present study (TCbio 19-704). All participants provided written informed consent in line with the Declaration of Helsinki.

## Results

### Patient characteristics

Longitudinal PBMC and serum samples collected at baseline, at ±3 weeks (Timepoint 2) and at ±6 weeks or upon irAEs before immunosuppression (Timepoint 3/Toxicity) from 44 ICI-treated patients (11 anti-PD-1 monotherapy with clinically relevant irAEs [TOX], 13 anti-PD-1 monotherapy without clinically relevant irAEs [NOTx], 10 combined ICI [cICI] TOX and 10 without TOX [cICI NOTx]) were included, along with 10 healthy donors (HD) (**Fig. 1, Table 1, Supplementary Table 3**). As expected for cICI TOX, these patients had more concomitant, earlier-onset and more severe irAEs than anti-PD-1 TOX patients (**Table 1**). Baseline absolute eosinophil count, monocyte-to-lymphocyte and neutrophil-to-lymphocyte ratios derived from complete blood counts were equal between TOX and NOTx patients, as well as T cell-to-monocyte ratio assessed by flow cytometry.

**Table 1.** Characteristics of patients and healthy donors

|  | Anti-PD-1 NOTx<br>(N=13) | Anti-PD-1 TOX<br>(N=11) | cICI NOTx<br>(N=10) | cICI TOX<br>(N=10) | Healthy donor<br>(N=10) | P value |
| --- | --- | --- | --- | --- | --- | --- |
| Age (yr)<br>Median (IQR) | 63.0 (57-69) | 73 (66.5-76.5) | 71.0 (58.25-75) | 49.5 (45.5-62.75) | 56.0 (38.75-60.75) | <0.01 |
| Sex (male), N (%) | 9 (69.2) | 8 (72.7) | 10 (100) | 8 (80.0) | 3 (30.0) | 0.012 |
| Primary tumor, N (%) |  |  |  |  |  |  |
| Melanoma | 9 (69.2) | 9 (81.8) | 3 (30.0) | 8 (80.0) | N/A | <0.01 |
| Non-small cell lung cancer | 0 (0.0) | 1 (9.1) | 0 (0.0) | 0 (0.0) |  |  |
| Renal cell carcinoma | 1 (7.7) | 0 (0.0) | 6 (60.0) | 2 (20.0) |  |  |
| Urothelial cell carcinoma | 3 (23.1) | 0 (0.0) | 0 (0.0) | 0 (0.0) |  |  |
| Other | 0 (0.0) | 1 (9.1)<sup>a</sup> | 1 (10.0)<sup>b</sup> | 0 (0.0) |  |  |
| Tumor stage, N (%) |  |  |  |  |  |  |
| III | 4 (30.8) | 4 (36.4) | 0 (0.0) | 0 (0.0) | N/A | 0.031 |
| IV | 9 (69.2) | 7 (63.6) | 10 (100.0) | 100 (100.0) |  |  |
| Time-to-toxicity (weeks)<br>Median (IQR) | N/A | 8.0 (6-12) | N/A | 4.5 (3-6) | N/A | 0.076 |
| Number of toxicities |  |  |  |  |  |  |
| 1 | N/A | 9 (81.8) | N/A | 5 (50.0) | N/A | 0.28 |
| 2 |  | 2 (18.2) |  | 2 (20.0) |  |  |
| 3 |  | 0 (0.0) |  | 2 (20.0) |  |  |
| >3 |  | 0 (0.0) |  | 1 (10.0) |  |  |
| Highest grade (CTCAE v5) |  |  |  |  |  |  |
| II | N/A | 8 (72.7) | N/A | 0 (0.0) | N/A | <0.01 |
| III |  | 2 (18.2) |  | 8 (80.0) |  |  |
| IV |  | 1 (9.1) |  | 2 (20.0) |  |  |
| Preexistent autoimmune<br>disease, N (%) | 0 (0.0) | 0 (0.0) | 1 (10.0) | 0 (0.0) | 0 (0.0) | 0.45 |

<sup>a</sup>Squamous cell carcinoma of unknown primary, <sup>b</sup>Mismatch repair deficient colorectal cancer. Groups are compared by Kruskal-Wallis (continuous variables) or Fisher's exact test (categorical variables). Abbreviations: anti-PD-1 denotes 'anti-PD-1 monotherapy', cICI 'combined immune checkpoint inhibition', CTCAE 'Common Terminology Criteria for Adverse Events', IQR 'interquartile range', N/A 'not applicable', NOTx 'no clinically relevant irAEs', TOX 'with clinically relevant irAEs', yr 'years'.

**Figure 1.**
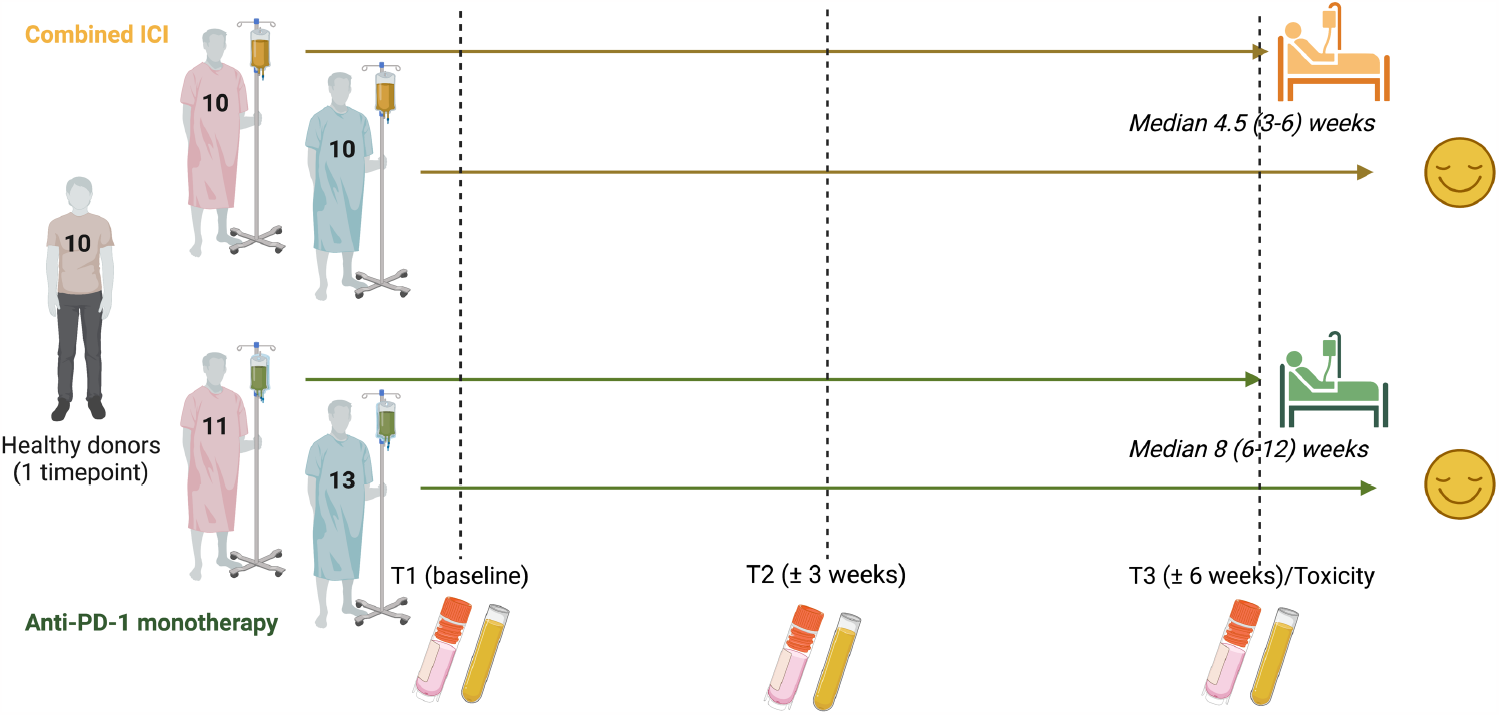
Design of multicolor longitudinal flow cytometry study. Peripheral blood mononuclear cell (PBMC) and serum samples from immune checkpoint inhibitor (ICI)-treated patients developing or remaining free of clinically relevant immune-related adverse events (irAEs) were included at baseline, ±3 weeks into treatment and at ±6 weeks or upon irAE onset (for which median time-to-onset and inter-quartile range are shown). Healthy donor PBMCs and serum were included for comparison. Created with BioRender.com.

### CD4_EM_ T cell proliferation is strongly associated with irAEs in combined ICI

We extensively characterized the dynamics of peripheral T and B cell immunity during ICI treatment and towards the manifestation of toxicity using multicolor flow cytometry. We explored shared features among subjects, with principal component analysis (PCA) with all 159 readout parameters from panels in **Supplementary Table 1** pooled together. cICI TOX patients, especially on-treatment timepoints, partially separated from all other subjects, including healthy donors, mainly through principal component 2 (**Fig. 2a, Supplementary Table 4**).

**Figure 2.**
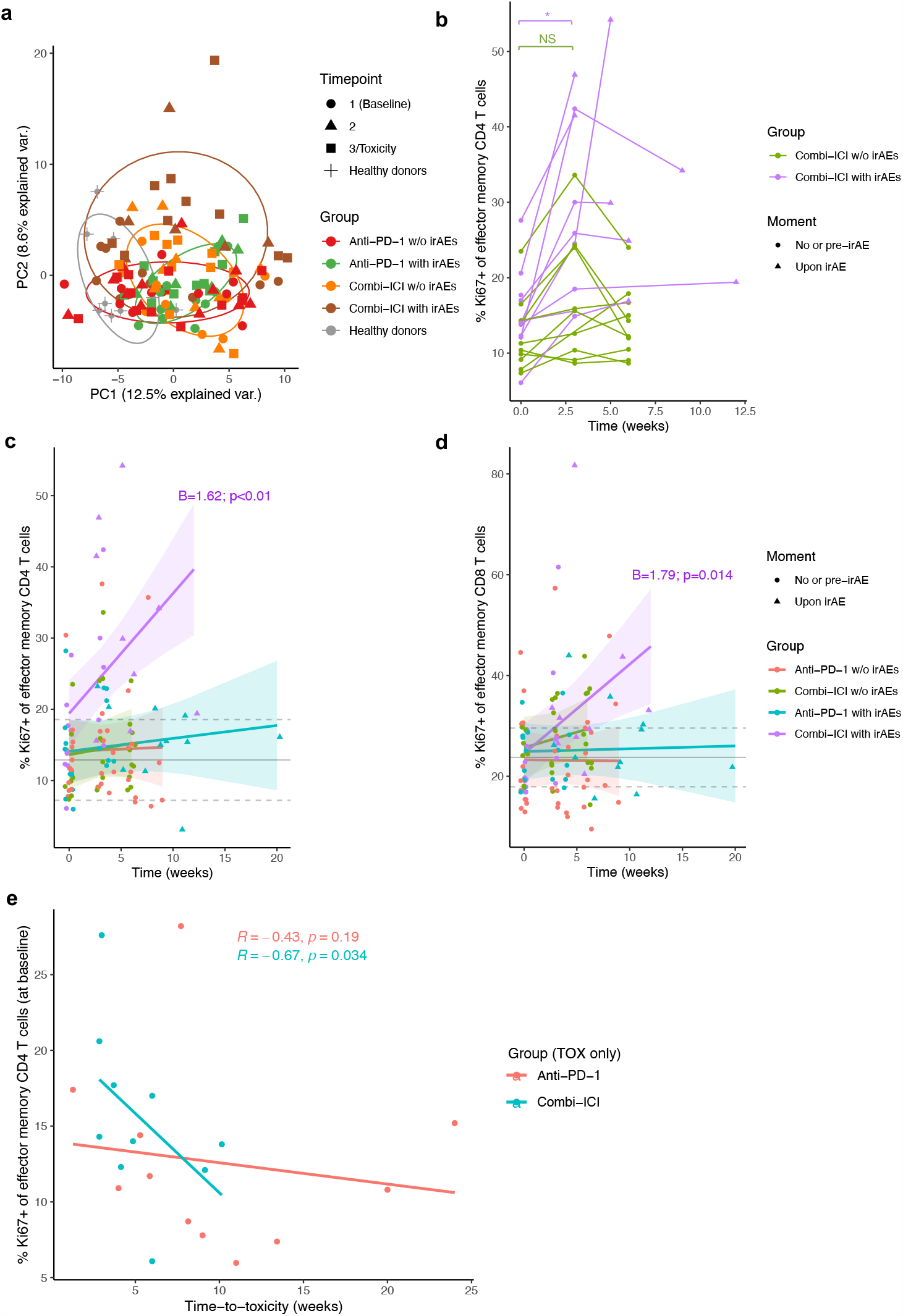
Effector memory CD4^+^ T cell proliferation increases over time in combined-ICI treated patients with toxicity, relative to other groups, and is at baseline associated with early irAE onset. (**a**) Plot showing clustering of different groups in principal component analysis (PCA) with pooled flow cytometric data including all 159 readout parameters across panels. (**b**) Individual patient trajectories of the percentage Ki67^+^ of CD4^+^ effector memory (CD4_EM_) T cells. Comparison by unpaired Wilcoxon test because of missing paired data for some patients. (**c**,**d**) Percentage proliferating of total CD4_EM_ (**c**) and CD8_EM_ (**d**) T cells over time. Only significant coefficients from mixed-effects models for the interaction term with time (‘B’), indicating statistically significant change compared to other groups, are shown. Gray solid and dashed lines indicate mean healthy donor level with 95% confidence interval. (**e**) Correlation between baseline CD_EM_ proliferation rate and time-to-toxicity, stratified by ICI-regimen (by Spearman correlation). irAE: immune-related adverse event, NS: not significant, w/o: without, * *P*<0.05

Based on the expression of CD27 and CD45RA, distributions of CD4^+^ and CD8^+^ T effector memory (EM, CD27^−^CD45RA^−^), central memory (CM, CD27^+^CD45RA^−^), terminally differentiated effector memory re-expressing CD45RA (EMRA, CD27^−^CD45RA^+^) and naive (CD27^+^CD45RA^+^) subsets were analyzed. Although pre-treatment CD4_EM_ frequency has been reported as irAE predictor [10], baseline proportions of CD4^+^ and CD8^+^ T subsets were similar between TOX and NOTx patients (**Supplementary Fig. 1a**). Compared to ICI-treated patients, healthy donors had fewer CD4_EM_ T cells and more naive CD8^+^ T cells (**Supplementary Fig. 1a**). During ICI treatment, and even upon onset of irAEs, relative frequencies of naive/memory subsets remained unchanged, except for an increase in CD8_EM_ T cells in cICI TOX (**Supplementary Fig. 1b**). However, group-specific differences in proliferation (based on Ki67) over time were present. A significant increase in CD4_EM_ proliferation from baseline to Timepoint 2 was observed in all cICI TOX patients, but not in cICI NOTx patients (**Fig. 2b**). To enable comparison of all five groups (TOX or NOTx in anti-PD-1 monotherapy or cICI and healthy donors) simultaneously and over time, we graphically summarized patient-level data by plotting group-specific linear mixed models. cICI TOX, but not NOTx or anti-PD-1-treated patients showed significantly increased proliferation in CD4_EM_, CD4_CM_, CD8_EM_, CD8_CM_ and CD8_EMRA_ T cells (**Fig. 2c,d**, **Supplementary Fig. 1c-e**).

While the latter effects pertain to the (early) effects of ICI treatment, we were also interested in baseline parameters associated with irAE development. Higher CD4_EM_ proliferation before the first ICI administration was associated with shorter time-to-toxicity (ρ = -0.53, *P* = 0.013), but this association was entirely driven by the cICI group (ρ = -0.67, *P* = 0.034, **Fig. 2e**). In most cICI treated TOX patients, the increase in Ki67 expression reached a plateau at 3 weeks after ICI initiation (**Fig. 2b**). Both cICI TOX patients with longest time-to-toxicity developed hypophysitis, a presumably complement-mediated irAE with distinct pathogenesis from most other toxicities [26]. This further strengthens the association of baseline CD4_EM_ proliferation with early-onset toxicity after cICI.

### Th1- and Th17-associated immune responses primarily delineate irAEs after combined ICI, while anti-PD-1 TOX features only a modest Th1-associated response

Expanding further on the CD4^+^ compartment, we investigated immune response skewing by the percentages of CXCR3^+^CCR4^-^, IFN-γ^+^ and IL-17^+^ CD4^+^ T cells. Relative abundance of all three subsets remained constant over time and neither correlated with ICI regimen, nor with toxicity development (data not shown). Although we found no differences in the percentage of IFN-γ^+^ CD4^+^ T cells over time (**Fig. 3a**), cycling CXCR3^+^CCR4^-^ (Th1-associated) CD45RO^+^ memory CD4^+^ T cells were dominant in cICI TOX relative to all other groups (**Fig. 3b**). Correspondingly, we observed increased proliferation of CXCR3^+^ CD45RO^+^ memory CD8^+^ T cells in cICI TOX patients (**Supplementary Fig. 2a**). These findings were corroborated by serum multiplex data, which showed increased serum levels of Th1-associated cytokines IL-12, TNF-α and IFN-γ in cICI TOX, and CXCL9 and CXCL10 both in anti-PD-1 and cICI TOX upon Timepoint 3/Toxicity (**Fig. 3c,d**, **Supplementary Fig. 2b**–**e**).

**Figure 3.**
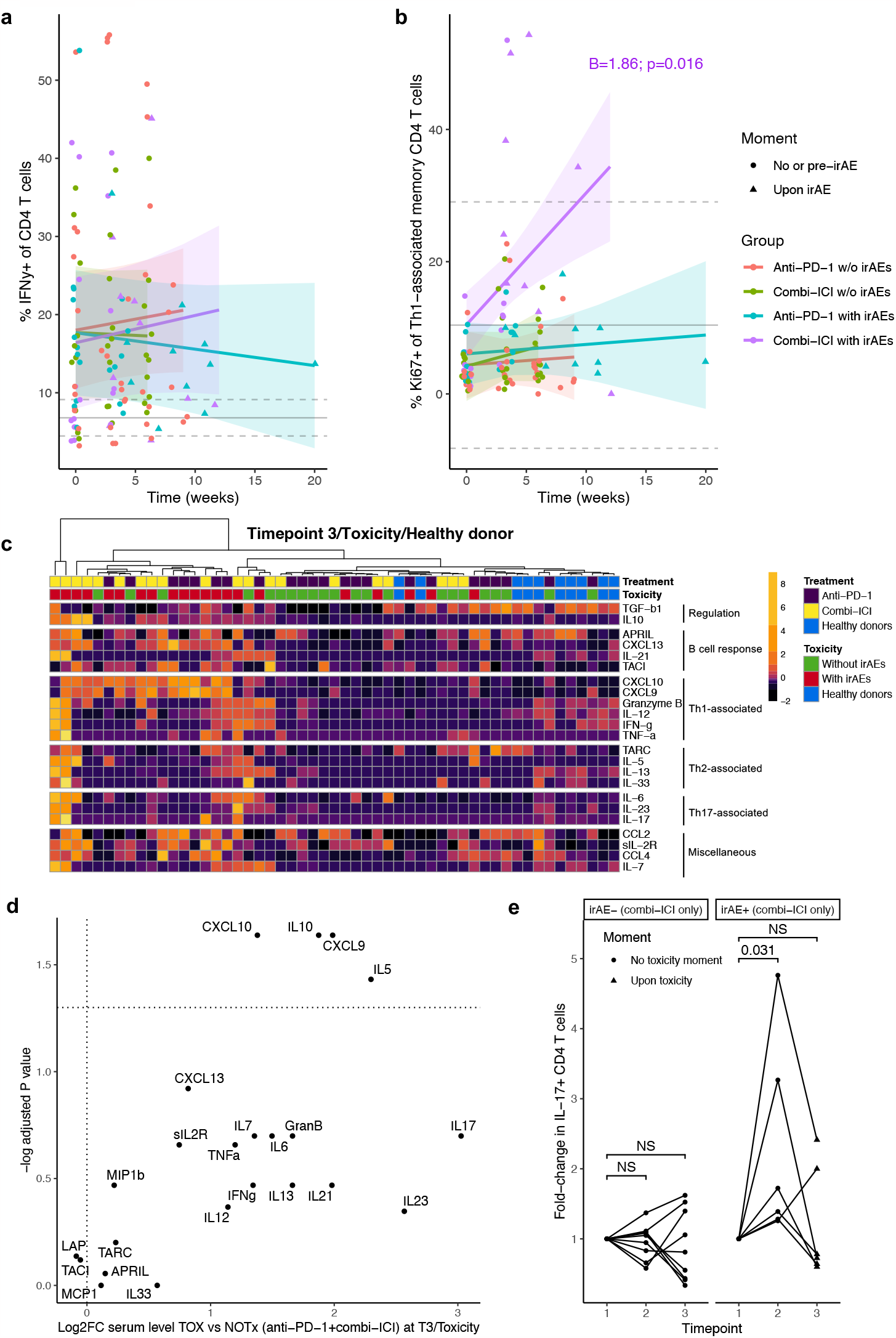
Both Th1- and Th17-associated immune responses are observed in combined-ICI treated patients with toxicity, whereas only enhanced Th1-associated immunity is associated with anti-PD-1 associated toxicity. (**a**,**b**) Percentage IFN-α^+^ of total CD4^+^ T cells (**a**) and percentage proliferating of CXCR3^+^CCR4^-^ Th1-associated CD4^+^ T cells (**b**) over time. Only significant coefficients from mixed-effects models for the interaction term with time (‘B’), indicating statistically significant change compared to other groups, are shown. Gray solid and dashed lines indicate mean healthy donor level with 95% confidence interval. (**c**) Heatmap showing serum levels of cytokines and chemokines after ±6 weeks (T3), upon irAE onset (Toxicity) or in healthy donors. Data are scaled by individual proteins across timepoints (see **Supplementary Fig. 3b**,**c**), enabling direct comparison of individual proteins between three timepoints. Unsupervised clustering of patients by Manhattan distance. (**d**) Volcano plot showing relative increase in serum protein levels at Timepoint 3/Toxicity in all patients with irAEs (TOX; anti-PD-1 and cICI combined) versus all patients without irAEs (NOTx). Full protein names are in **Supplementary Table 2**. (**e**) Fold-change in percentage IL-17^+^ of CD4^+^ T cells relative to baseline in cICI-treated patients without irAEs (left; NOTx) and with (right; TOX) irAEs. irAE: immune-related adverse event, NS: not significant, w/o: without.

We further characterized the kinetics of IL-17 producing CD4^+^ T cells, which is especially relevant since inhibition of Th17 differentiation by the IL-6 receptor blocker (IL-6RB) tocilizumab is clinically being tested to uncouple ICI toxicity from efficacy [27]. After the first treatment cycle, cICI TOX, but not anti-PD-1 TOX or NOTx patients demonstrated a vast increase in IL-17^+^ T cells (**Fig. 3e, Supplementary Fig. 2f**). Ki67 expression in IL-17^+^ CD4^+^ T cells was non-significantly higher in TOX compared to NOTx across all timepoints, including baseline, regardless ICI regimen (**Supplementary Fig. 2g**). The relative decrease of IL-17^+^ T cells from timepoint 2 to irAE onset in cICI TOX is likely due to relative expansion of other subsets, as serum concentrations of Th17-associated cytokines IL-6, IL-23 and IL-17 remained stable or even increased from Timepoint 2 towards irAE onset (**Fig. 3c, Supplementary Fig. 2c**) while IL-23 and IL-17 even show a greater than eightfold increase in cICI TOX compared to cICI NOTx patients at Timepoint 3/Toxicity (**Supplementary Fig. 2e**).

Together, our data confirm that cICI TOX is largely Th1/Th17 driven, while anti-PD-1 TOX only features modest Th1 enhancement relative to anti-PD-1 NOTx as assessed by serum multiplex data. Furthermore, these human data suggest that IL-6RB therapy to mitigate ICI toxicity may be more effective in cICI than in anti-PD-1 monotherapy, although preclinical models have demonstrated its efficacy both in anti-CTLA-4 and anti-PD-1 monotherapies [27].

### PD-1^+^LAG-3^+^ CD8^+^ T cells bear intact inflammatory and cytotoxic potential regardless development of irAEs

Next, we investigated the cytotoxic potential of CD4^+^ and CD8^+^ T cells in relation to co-inhibitory receptor expression. We used CD95 expression to select for the antigen-experienced CD8^+^ T cell pool [28], which largely overlaps with CD45RO^+^ memory cells but also includes T_EMRA_s (**Fig. 4a**). First, we created t-SNE and FlowSOM visualizations of CD3^+^ T cells from Timepoint 3/Toxicity across all patients and healthy donors, to explore the variety of subsets in an unbiased way. Proinflammatory cells with high potential for IFN-γ production were represented mainly across the CD8^+^ compartment (**Fig. 4b**). Within the CD8^+^ compartment, granzyme B production was especially high in the subset expressing senescence marker CD57 (**Fig. 4b**). Although this subset appeared brighter in the anti-PD-1 and cICI TOX density plots (**Fig. 4b**), we found no statistical difference in CD57^+^ CD8^+^ T cell abundance between groups at Timepoint 3/Toxicity. Labels assigned based on manual gating matched unsupervised clustering results, demonstrating that our gating strategy adequately covers relevant biological variation (**Supplementary Fig. 3a**).

**Figure 4.**
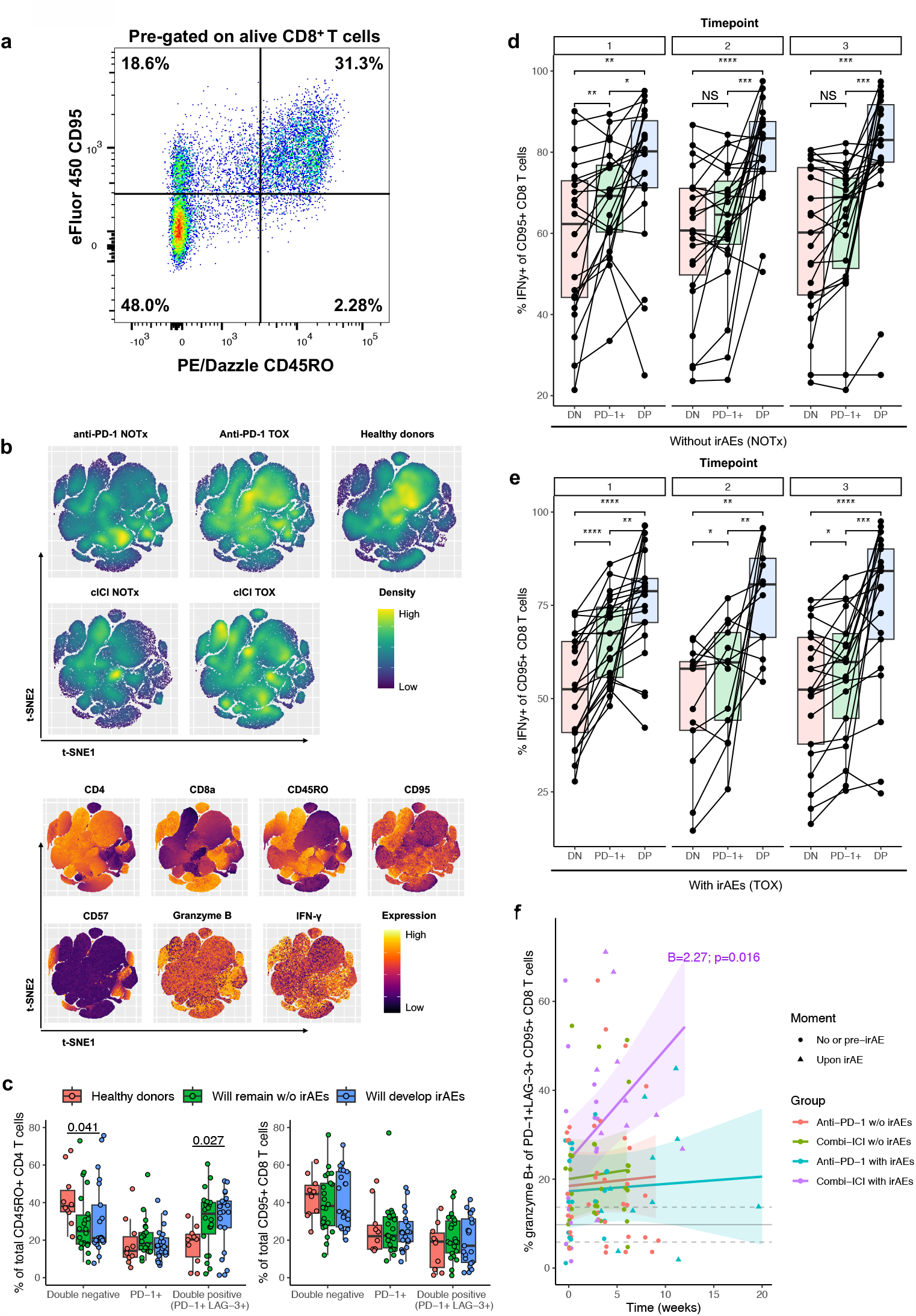
PD-1^+^LAG-3^+^ double positive CD8^+^ memory T cells retain their inflammatory potential regardless the development of toxicity. (**a**) Representative flow cytometry plot of CD95 and CD45RO expression in CD8^+^ T cells. (**b**) t-SNE visualizations for a random 10% subset of all CD3^+^ T cells from all patients at Timepoint 3/Toxicity and healthy donors, with relative abundance shown in density plots (top) and expression of lineage markers, IFN-γ and granzyme B in overlay plots (bottom). (**c**) Baseline percentages of CD45RO^+^ CD4^+^ (left) and CD95^+^ CD8^+^ (right) T cells positive for PD-1 alone or PD-1 and LAG-3. Comparisons by Kruskal-Wallis tests. (**d**,**e**) Percentages of CD95^+^CD8^+^ T cells producing IFN-γ after PMA/ionomycin stimulation, depending on inhibitory receptor expression pattern, in patients without (**d**) or with irAEs (**e**) stratified by time of sample collection; anti-PD-1- and cICI-treated patients are combined. Comparisons by pairwise paired Wilcoxon tests with Benjamini-Hochberg correction for multiple testing. (**f**) Percentage of granzyme B producing PD-1^+^LAG-3^+^ CD95^+^CD8^+^ T cells over time. Significant coefficient (interaction term with time; ‘B’) from mixed-effects model is shown. Gray solid and dashed lines indicate mean healthy donor level with 95% confidence interval. cICI: combined ICI, irAE: immune-related adverse event, NS: not significant, w/o: without. * *P*<0.05, ** *P*<0.01, *** *P*<0.001, **** *P*<0.0001.

Subsequentially, we investigated co-expression patterns of classical exhaustion markers PD-1 and LAG-3. At baseline, PD-1^+^LAG-3^+^ double-positive (DP) CD8^+^ T cells were least prevalent across groups, while DP CD4^+^ T cells were higher and made up the largest fraction of CD4^+^ T cells in both TOX and NOTx patients, compared to healthy donors (**Fig. 4c**). We found no differences in expression patterns of PD-1 and LAG-3 occurring between TOX and NOTx post-treatment (**Supplementary Fig. 3b**).

Since merely expression of inhibitory receptors is not indicative of differentiation or functional status, we assessed proinflammatory and cytotoxic capacity by measurement of IFN-γ and granzyme B production after restimulation with PMA and ionomycin. Compared to double-negative (DN) T cells, a much larger proportion of DP (and to a lesser extent PD-1^+^LAG-3^-^) CD8^+^ T cells produced IFN-γ (**Fig. 4d,e**). This indicates preserved proinflammatory potential of DP cells and is supported by transcriptomic data in circulating PD-1^+^TIGIT^+^ CD8^+^ T cells, mostly co-expressing LAG-3, in melanoma and Merkel cell carcinoma [29]. In contrast, we found that granzyme B production was drastically lower in DP relative to DN CD8^+^ T cells, regardless timepoint or the development of irAEs (**Supplementary Fig. 3c**,**d**). Still, cICI TOX featured a profound increase in CD8^+^ granzyme B production over time relative to other groups, especially in the DP subset (**Fig. 4f, Supplementary Fig. 3e**,**f**).

In conclusion, our data suggest remaining cytokine production potential in supposedly senescent CD57^+^ CD8^+^ T cells. Moreover, PD-1^+^LAG-3^+^ DP CD8^+^ T cells retain their proinflammatory and cytotoxic potential, while towards irAEs after cICI the fraction of granzyme B producing cells increases especially in this DP subset.

### No changes in B cell subsets are observed, but serum cytokine levels indicate germinal center activation in combined ICI TOX

Then we set out to analyze the CD19^+^ B cell compartment over time. Although early increase in CD21^lo^ B cells (mainly representing a memory subset) has been associated with irAEs following cICI [11, 30], we found no changes in CD21^lo^ B cells following ICI, while CD27^+^ memory B cells in cICI TOX even decreased (**Fig. 5a,b**). Moreover, we observed no B cell activation, measured by CCR6 expression (**Supplementary Fig. 4a**). The fraction of IgM^-^ class-switched B cells also remained unchanged over time in all groups (**Supplementary Fig. 4b**). Despite lacking indications for B cell activation and maturation based on flow cytometry data, increasing serum levels of APRIL, CXCL13 and IL-21 towards toxicity suggest germinal center activity or tertiary lymphoid structure (TLS) formation, again especially in cICI TOX (**Fig. 5c, Fig. 3c, Supplementary Fig. 2b**,**c**). Besides, higher CD27^+^CD38^hi^ plasmablast abundance at baseline was associated with shorter time-to-toxicity in cICI TOX (**Fig. 5d**). In sum, these data support a role for adaptive humoral immunity following cICI that is more pronounced in TOX than NOTx, although no changes of B cell abundances were observed in peripheral blood.

**Figure 5.**
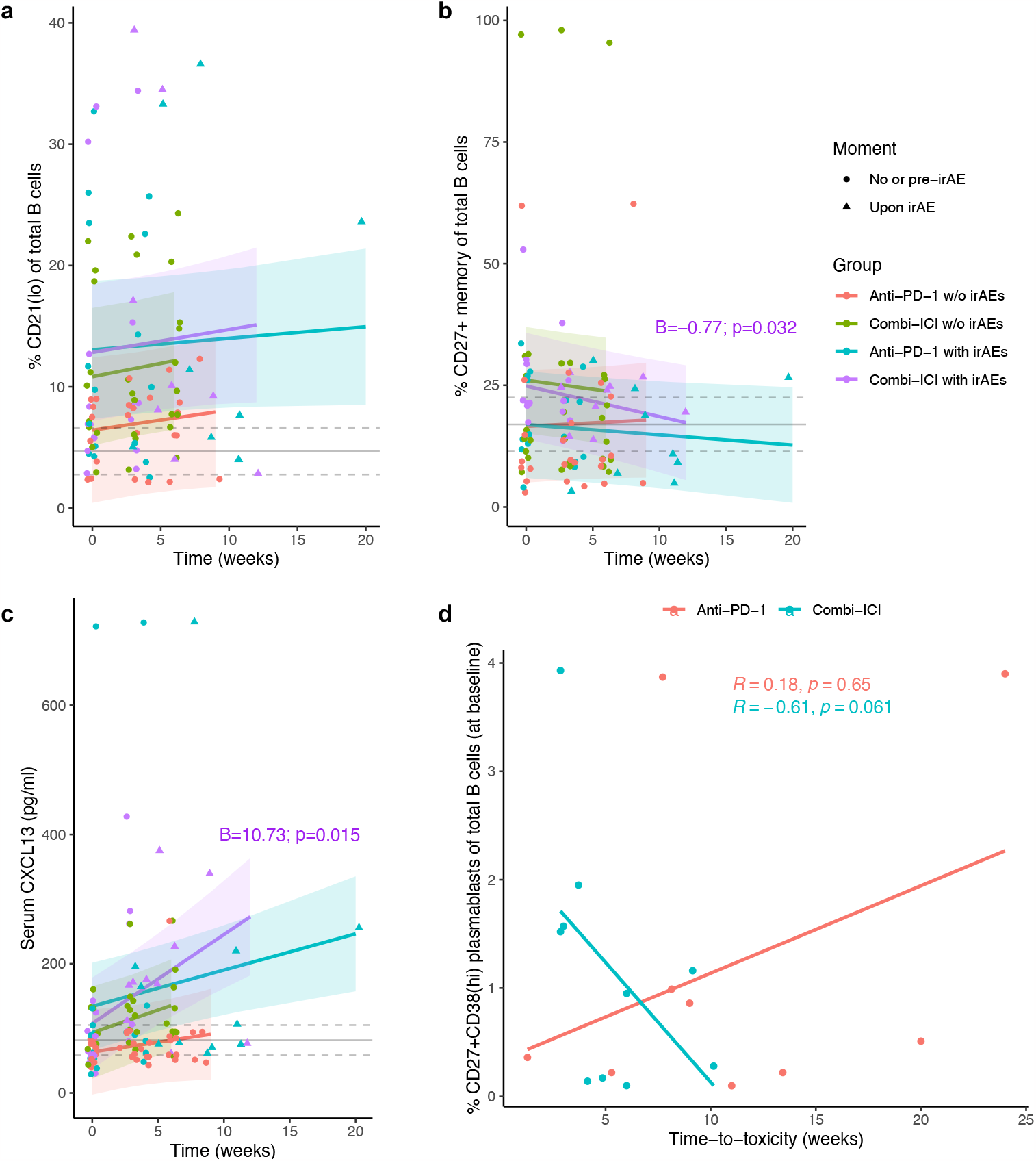
Increasing serum CXCL13 levels and higher baseline plasmablast abundance in patients with early toxicity suggest a role for adaptive humoral immunity in toxicity after combined ICI. (**a**-**c**) Percentages of CD21^lo^ (**a**) and CD27^+^ memory (**b**) B cells of total B cells and serum level (in pg/ml) of CXCL13 (**c**) over time. Only significant coefficients (interaction terms with time; ‘B’) from mixed-effects models are shown. Gray solid and dashed lines indicate mean healthy donor level with 95% confidence interval. (**d**) Correlation between baseline percentage CD27^+^CD38^hi^ plasmablasts of total B cells and time-to-toxicity, stratified by ICI-regimen (by Spearman correlation). irAE: immune-related adverse event, w/o: without.

### Regulatory activity by Tregs is preserved upon systemic inflammation in the context of irAEs

Finally, we sought to characterize the response of regulatory T (Treg) cells after ICI treatment. Abundance of forkhead box protein 3 (FOXP3)^+^ CD25^+^ CD4^+^ Tregs increased over time, especially in cICI TOX patients (**Fig. 6a**). Already at baseline, Tregs from patients showed higher expression of T-box transcription factor TBX21 (T-bet) compared with healthy donors (**Fig. 6b**). This may be indicative of a more activated Th1-polarized state, even before the introduction of ICI. To assess whether Tregs can adequately respond to systemic inflammation inflicted by ICI, we compared expression levels of co-inhibitory and co-stimulatory receptors. At Timepoint 3/Toxicity, mainly cICI TOX patients clustered together, marked by upregulation of CTLA-4, ICOS and TIGIT (**Fig. 6c**). Notably, the only two cICI TOX patients with grade 4 irAEs clustered distinctly, characterized by Tregs that failed to upregulate CTLA-4 or TIGIT. In sum, these findings indicate that peripheral Treg function upon systemic inflammation is generally preserved after ICI, while Treg dysfunction may contribute to more severe irAEs.

**Figure 6.**
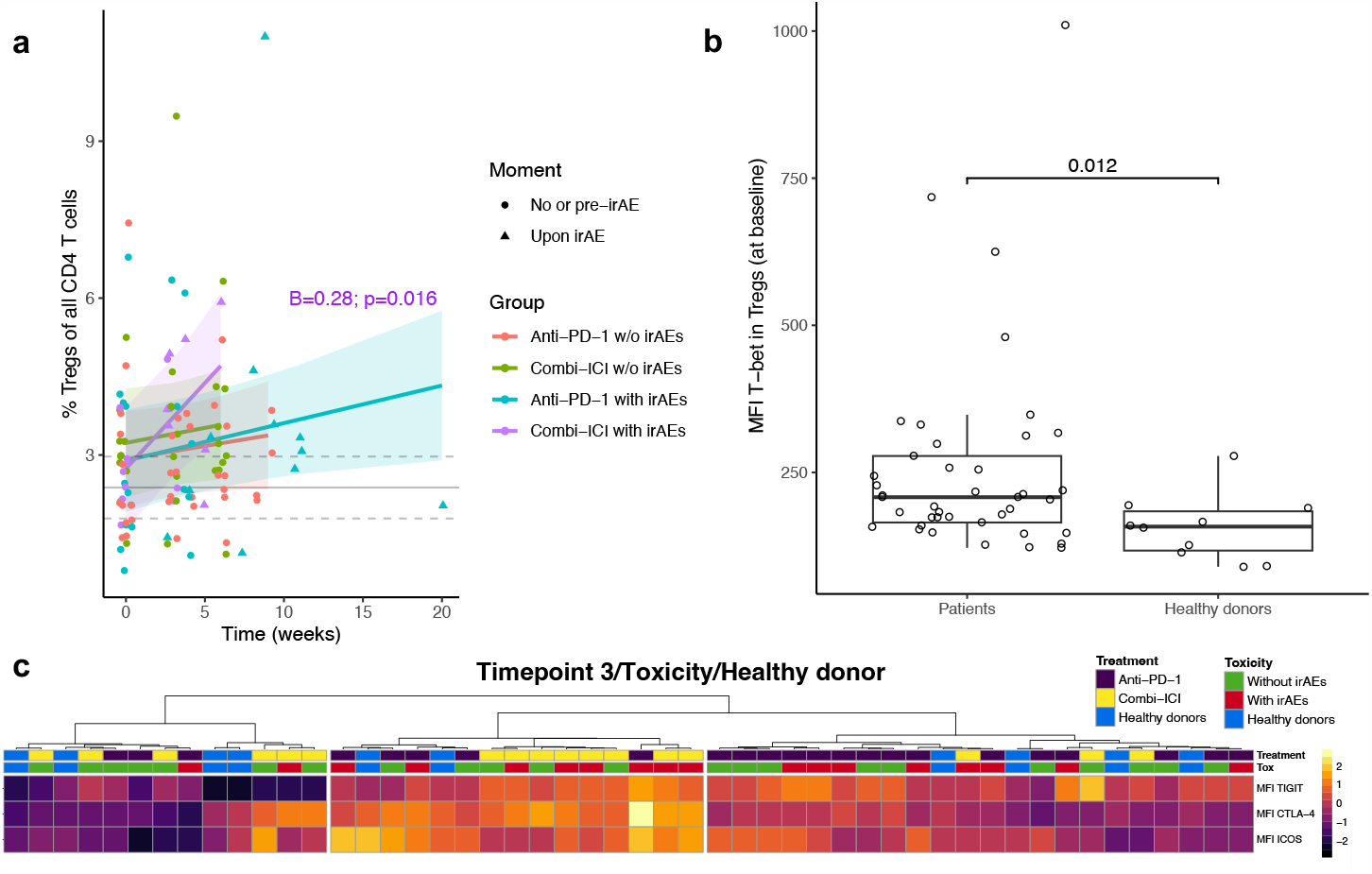
Peripheral regulatory T cell abundance and function are generally preserved upon toxicity. (**a**) Percentage of CD25^+^FOXP3^+^ CD4^+^ Treg cells of total CD4^+^ T cells over time. Significant coefficient (interaction term with time; ‘B’) from mixed-effects model is shown. Gray solid and dashed lines indicate mean healthy donor level with 95% confidence interval. (**b**) Comparison of median fluorescence intensity (MFI) of T-bet in Tregs at baseline between healthy donors and all ICI-treated patients, by Wilcoxon test. (**c**) Expression levels (by MFI) of inhibitory and costimulatory receptors in Tregs (for patients Timepoint 3/Toxicity), hierarchically clustered by Euclidean distance. irAE: immune-related adverse event, w/o: without.

## Discussion

Longitudinal studies into mechanisms underlying irAEs can importantly contribute to establishing evidence-based treatment strategies. Therefore, we studied lymphocyte dynamics in peripheral blood of healthy donors and ICI-treated patients with and without irAEs from baseline to several weeks into treatment. Key findings are that cycling Th1-associated effector memory CD4^+^ T cells appear important drivers of acute irAEs, specifically in cICI. Besides, Th17- and Th2-associated responses contribute to the development of irAEs following cICI, but no changes in abundance or activation of B cell subsets were found. In contrast, anti-PD-1-associated irAEs were characterized by a modestly enhanced Th1 response compared to anti-PD-1-treated patients without irAEs. This likely relates to the less acute nature of anti-PD-1 associated toxicity. We showed that peripheral blood PD-1^+^LAG-3^+^ CD8^+^ T cells retained their proinflammatory potential and that especially within this DP subset the fraction of granzyme B producing cells increases towards cICI TOX relative to other patient groups.

Our key findings are corroborated by a recent study which showed that irAEs are associated with early proliferation of peripheral blood (activated) CD4^+^ and CD8^+^ T cell subsets in a Th1-skewed milieu (based on serum CXCL9/10/11 and IFN-α) [9]. In contrast to our findings, this group also found a cellular Th1-associated response for irAEs after anti-PD-1. This may be due to the difference in sampling of the first on-treatment timepoint at 7-14 days in [9] versus ±3 weeks in our study. Indeed, Nuñez et al. found that at later timepoints the percentage activated CD4^+^ and CD8^+^ of all T cells had returned to baseline levels in patients with irAEs [9]. This also fits the observation that cycling CD8^+^ T cells peak at seven days after anti-PD-1 treatment before they gradually return towards baseline levels three weeks post-treatment [31], possibly because of migration of proliferating cells into tissues.

ICI may induce proliferation and reinvigorate antitumor responses in different non-terminally exhausted cell populations, including PD-1^+^ CD8^+^ T cells [32, 33]. We showed that PD-1^+^LAG-3^+^ (DP) T cells produced highest amounts of IFN-α after restimulation, which may indicate intact proinflammatory potential *in vivo*. However, phenotypically similar T cells may exert differential function dependent on location [34]. Besides, despite T cell transcriptomic overlap of proinflammatory genes and exhaustion markers [35, 36], *ifng* translation may be hampered in dysfunctional T cells [37]. In addition to IFN-α production, cICI TOX featured a profound increase in granzyme B production over time, especially in DP CD8^+^ T cells, which could indicate reinvigoration of dysfunctional cells and *de novo* activation. In sum, our study presents DP T cells as most potent proinflammatory and cytotoxic cells, but to what extent this potential is deployed *in vivo* likely depends on ICI regimen.

We observed ongoing proliferation in the CD8^+^ and, most prominently, CD4^+^ memory pools in cICI TOX. Our results are consistent with other studies indicating increased CD4^+^ and CD8^+^ clonal expansion and repertoire diversification in patients with irAEs [10, 15, 38-40]. Moreover, CD4_EM_ T cell expansion in patients with irAEs parallels CD4 memory abundance in classical autoimmune disease [10]. Together, these data reinforce the hypothesis that cross- or autoreactive CD4_EM_ clones may be unleashed upon ICI treatment in some, especially early, irAEs.

Helper T cells may also provide help to autoantibody-producing B cells. In line with an antibody-mediated etiology, our serum cytokine data (including CXCL13 and IL-21) suggest germinal center activation or TLS formation both in cICI and anti-PD-1 monotherapy irAEs. CD4_EM_ T cells have been shown to contribute to thyroiditis development in anti-PD-1 treated mice with preexisting autoimmunity following anti-thyroglobulin immunization [13]. Recent work by Johannet et al. showed that baseline serum autoantibody signatures are predictive of severe irAEs (not limited to thyroiditis) with adjuvant anti-PD-1 and/or anti-CTLA-4 [41], which presents further evidence that preexistent subclinical autoimmunity may be at the root of various irAEs. Importantly, CXCL13 increase does not *per se* indicate TLS formation, as tumor-reactive T cells identified through *CXCL13* expression can also be a source that expands upon treatment in ICI-responders [42].

The role of B cells in irAE development is further solidified by human data on increasing CD21^lo^ B cells in association with irAEs in the setting of cICI [11, 30]. We were not able to reproduce these findings, which may be due to different timing in blood collection and B cells migrating into peripheral tissues shortly following activation. A recent study found that aged anti-PD-1-treated mice developed TLS-like aggregates in irAE-affected tissue in a CXCL13-dependent fashion and that B cell depletion could attenuate ICI-inflicted toxicity [43]. So far, the use of rituximab for irAE management is limited to case reports [44]. These data together with our work reinforce B cells as a potential target for irAE treatment, although caution is warranted as TLS activation has also been strongly associated with clinical response to ICI [45].

Strengths of our study include the longitudinal design with multiple on-treatment timepoints, the stratified analysis for anti-PD-1 and combined ICI treatment and the variety of measured T and B cell parameters. This allowed investigation of very early treatment-specific immunological effects that precede irAE onset. Our study has several limitations. First, age and sex were unevenly distributed among groups. Second, there was considerable heterogeneity in type of irAEs, also within different strata. However, this simultaneously presents as a strength: we clearly found significant and clinically relevant differences between groups that would be less generalizable to the general irAE patient population if obtained within a controlled and irAE-homogenized cohort. Third, our study was powered to discover irAE-related differences stratified by ICI regimen. Therefore, it did not allow for further stratification by ICI response, which would moreover require restriction to a single tumor type. Lastly, our research was limited to the peripheral blood compartment.

In conclusion, we show that the peripheral blood immune response substantially differs between patients treated with cICI and anti-PD-1 monotherapy, and between cICI-treated patients with and without irAEs. Toxicity is clearly dominated by enhanced Th1-associated immunity, but we showed that the Th17 pathway and germinal center activation also contribute to cICI toxicity. In this way, our study provides rationale to guide irAE treatment schemes based on ICI regimen and to deploy specific T-cell-directed (second-line) strategies, such as anti-IL-6R, especially in cICI-associated irAEs.

## Data Availability

Data and scripts for statistical analysis are available upon reasonable request.

## Statements & Declarations

### Author contributions

Conceptualization: FW, KPMS, MJME. Project administration and Resources: RJV, MJME, ASRL, KPMS. Data curation: MJME, SAW, RJV. Formal Analysis: MJME, FW, KPMS. Writing - original draft: MJME. Writing – review and editing: all authors. Supervision: FW, KPMS.

### Data availability

Data and scripts for statistical analysis are available upon reasonable request.

### Ethics approval

This study was approved by the University Medical Center Utrecht biobank committee (TCbio 18-123).

### Consent to participate

All participants provided written informed consent in line with the Declaration of Helsinki.

## Acknowledgements

We thank the patients, their families and caregivers, healthy donors, clinical staff and UNICIT consortium members. We would like to thank the Multiplex Core Facility of the University Medical Center Utrecht for performing the multiplex immunoassays.

