## Supplementary material for "Toxicity-specific peripheral blood T and B cell dynamics in anti-PD-1 and combined immune checkpoint inhibition"

### Supplementary Tables

#### Supplementary Table 1. Flow cytometry panels

| Antigen | Fluorophore | Clone | Company | Catalog number | Lot number(s) | Staining | Dilution |
|---|---|---|---|---|---|---|---|
| ***Panel 1 (B cells)*** | | | | | | | |
| IgG | FITC | polyclonal | Southern Biotech | 2042-02 | K2419-Z619E | Surface | 500 |
| CD38 | PerCP-Cy5.5 | HIT2 | BD | 551400 | 1159441 | Surface | 100 |
| CD27 | APC | L128 | BD | 337169 | 1105983 | Surface | 50 |
| CD3 | AF700 | UCHT1 | Biolegend | 300424 | B326013, B279942, B347088 | Surface | 50 |
| CD19 | APC-eF780 | HIB19 | eBioscience | 47-0199-42 | 2387508 | Surface | 20 |
| IgM | BV421 | G20-127 | BD | 562618 | 0328568 | Surface | 50 |
| Fixable viability dye | eFluor506 | - | Fisher Scientific | 15560607 | 2290923 | - | 1000 |
| CD21 | BV711 | B-ly4 | BD | 563163 | 1146348 | Surface | 20 |
| CD24 | PE-CF594 | ML5 | BD | 562405 | 1229319 | Surface | 200 |
| CCR6 | PE-Cy7 | R6H1 | eBioscience | 25-1969-42 | 2304422 | Surface | 100 |
| ***Panel 2 (T cell cytotoxicity)*** | | | | | | | |
| CD57 | FITC | HNK-1 | Biolegend | 359603 | B327318 | Surface | 100 |
| CD8α | PerCP-Cy5.5 | RPA-T8 | Biolegend | 301032 | B334771 | Surface | 1000 |
| CD3 | AF700 | UCHT1 | Biolegend | 300424 | B326013, B279942, B347088 | Surface | 50 |
| CD95 | eFluor450 | DX2 | eBioscience | 48-0959-42 | 2153486 | Surface | 100 |
| Fixable viability dye | eFluor506 | - | Fisher Scientific | 15560607 | 2290923 | - | 1000 |
| PD-1 | BV711 | EH12.1 | BD | 564017 | 1229849 | Surface | 100 |
| IgG4 (on-treatment) | biotin | HP6025 | Invitrogen | A10663 | 2309130, 2431369 | Surface | 50 |
| Streptavidin | BV711 | - | Biolegend | 405241 | B336306 | Surface | 100 |
| CD4 | BV785 | RPA-T4 | Biolegend | 300554 | B337916, B344396, B351096 | Surface | 50 |
| LAG-3 | PE | polyclonal | R&D | FAB2319P | AALE1619111 | Surface | 25 |
| CD45RO | PE-Dazzle | UCHL1 | Biolegend | 304247 | B301149 | Surface | 400 |
| Granzyme B | APC-Fire750 | QA16A02 | Biolegend | 372210 | B337650 | Intracellular | 50 |
| IFN-γ | PE-Cy7 | 4S.B3 | BD | 557844 | 1117951 | Intracellular | 200 |
| ***Panel 3 (T-helper subsets)*** | | | | | | | |
| CCR4 | FITC | 205410 | R&D | FAC1567F | LDG0621041 | Surface | 16.67 |
| CD8α | PerCP-Cy5.5 | RPA-T8 | Biolegend | 301032 | B334771 | Surface | 1000 |
| CD3 | AF700 | UCHT1 | Biolegend | 300424 | B326013, B279942, B347088 | Surface | 50 |
| Fixable viability dye | eFluor506 | - | Fisher Scientific | 15560607 | 2290923 | - | 1000 |
| CXCR3 | BV605 | G025H7 | Biolegend | 353728 | B329131, B339512, B349628 | Surface | 12.5 |
| CD45RA | BV711 | HI100 | Biolegend | 304138 | B334528 | Surface | 500 |
| CD4 | BV785 | RPA-T4 | Biolegend | 300554 | B337916, B344396, B351096 | Surface | 50 |
| CCR5 | PE | eBioT21/8 | eBioscience | 12-1957-42 | 2172554 | Surface | 50 |
| CD45RO | PE-Dazzle | UCHL1 | Biolegend | 304247 | B301149 | Surface | 400 |
| Ki67 | AF647 | B56 | BD | 558615 | 0342573, 1225152 | Intracellular | 50 |
| FOXP3 | eFLuor450 | PCH101 | eBioscience | 48-4776-42 | 2299833 | Intracellular | 50 |
| ***Panel 4 (naive/memory subsets)*** | | | | | | | |
| CD95 | FITC | DX2 | BD | 340479 | 1067724 | Surface | |
| CD8α | PerCP-Cy5.5 | RPA-T8 | Biolegend | 301032 | B334771 | Surface | 1000 |
| CD3 | AF700 | UCHT1 | Biolegend | 300424 | B326013, B279942, B347088 | Surface | 50 |
| CD27 | APC-eF780 | O323 | eBioscience | 47-0279-42 | 2241980, 2452256 | Surface | 20 |
| Fixable viability dye | eFluor506 | - | Fisher Scientific | 15560607 | 2290923 | - | 1000 |
| CD31 | BV605 | WM59 | BD | 562855 | 1099569, B283706 | Surface | 20 |
| CD45RA | BV711 | HI100 | Biolegend | 304138 | B334528 | Surface | 500 |
| CD4 | BV785 | RPA-T4 | Biolegend | 300554 | B337916, B344396, B351096 | Surface | 50 |
| Ki67 | AF647 | B56 | BD | 558615 | 0342573,1225152 | Intracellular | 50 |
| IL-2 | PB | MQ1-17H12 | Biolegend | 500324 | B317866 | Intracellular | 100 |
| IL-17A | PE | eBio64DEC17 | eBioscience | 12-7179-42 | 2331144 | Intracellular | 50 |
| IL-8 | PE-CF594 | G265-8 | BD | 563531 | 0311622 | Intracellular | 200 |
| ***Panel 5 (regulation)*** | | | | | | | |
| ICOS | FITC | C398.4A | Biolegend | 313506 | B311819 | Surface | 400 |
| TIGIT | PerCP-Cy5.5 | MBSA43 | eBioscience | 46-9500-42 | 2284173 | Surface | 50 |
| CD3 | AF700 | UCHT1 | Biolegend | 300424 | B326013, B279942, B347088 | Surface | 50 |
| CD45RA | APC-Cy7 | HI100 | Sony Biotech | 2120640 | 235800 | Surface | 50 |
| Fixable viability dye | eFluor506 | - | Fisher Scientific | 15560607 | 2290923 | - | 1000 |
| CD4 | BV785 | RPA-T4 | Biolegend | 300554 | B337916, B344396, B351096 | Surface | 50 |
| CCR8 | PE | L263G8 | Biolegend | 360603 | B334779 | Surface | 100 |
| CD25 | PE-Cy7 | M-A251 | BD | 557741 | 1068706 | Surface | 25 |
| CTLA-4 | APC | BNI3 | BD | 555855 | 0335549 | Intracellular | 12.5 |
| FOXP3 | eFLuor450 | PCH101 | eBioscience | 48-4776-42 | 2299833 | Intracellular | 50 |
| T-bet | PE-CF594 | O4-46 | BD | 562467 | 1209448 | Intracellular | 50 |
| ***Panel 6 (T cell-monocyte ratio)*** | | | | | | | |
| CD3 | PerCP-Cy5.5 | UCHT1 | Biolegend | 300430 | B331881 | Surface | 100 |
| CD14 | APC-eF780 | 61D3 | eBioscience | 47-0149-42 | 2284099 | Surface | 40 |

**Supplementary Table 2. Luminex analytes**

| Target | Full protein name |
|---|---|
| IL-5 | Interleukin-5 |
| IL-6 | Interleukin-6 |
| IL-7 | Interleukin-7 |
| IL-10 | Interleukin-10 |
| IL-12 | Interleukin-12 |
| IL-13 | Interleukin-13 |
| IL-17 | Interleukin-17 |
| IL-21 | Interleukin-21 |
| IL-23 | Interleukin-23 |
| IL-33 | Interleukin-33 |
| TNF-$\alpha$ | Tumor necrosis factor alpha |
| IFN-$\gamma$ | Interferon gamma |
| APRIL | A proliferation-inducing ligand |
| CCL2 (MCP1) | C-C motif ligand 2 (monocyte chemoattractant protein 1) |
| CCL4 (MIP-1$\beta$) | C-C motif ligand 4 (macrophage inflammatory protein) |
| CCL17 (TARC) | C-C motif ligand 17 (thymus- and activation-regulated chemokine) |
| CXCL9 | C-X-C motif ligand 9 |
| CXCL10 | C-X-C motif ligand 10 |
| CXCL13 | C-X-C motif ligand 13 |
| sILR2 | Soluble interleukin-2 receptor |
| GzmB | Granzyme B |
| TGF-$\beta$1 (LAP) | Transforming growth factor beta 1 (latency-associated peptide) |
| TACI | Transmembrane activator and CAML interactor |

**Supplementary Table 3. Extended characteristics of patients and healthy donors**

| | **Anti-PD-1 NOTx (N=13)** | **Anti-PD-1 TOX (N=11)** | **cICI NOTx (N=10)** | **cICI TOX (N=10)** | **Healthy donor (N=10)** | ***P* value** |
|---|---|---|---|---|---|---|
| Timepoint 2 available, N (%)[a] | 13 (100.0) | 7 (63.6) | 10 (100.0) | 6 (60.0) | N/A | **0.011** |
| Time-to-timepoint 2 (weeks) | | | | | | |
| Median (IQR) | 3.0 (3-4) | 4.0 (3.5-4) | 3.0 (3-3) | 3.0 (3-3) | N/A | **<0.01** |
| Time-to-timepoint 3 (weeks) | | | | | | |
| Median (IQR) | 6.0 (6-8) | N/A | 6.0 (6-6) | N/A | N/A | 0.11 |
| Best overall tumor response per RECIST1.1 | | | | | | |
| Not evaluable | 2 (15.4) | 3 (27.3) | 0 (0.0) | 2 (20.0) | N/A | 0.51 |
| Progressive disease | 6 (46.2) | 1 (9.1) | 5 (50.0) | 3 (30.0) | | |
| Stable disease | 2 (15.4) | 4 (36.4) | 1 (10.0) | 2 (20.0) | | |
| Partial response | 2 (15.4) | 2 (18.2) | 4 (40.0) | 2 (20.0) | | |
| Complete response | 1 (7.7) | 1 (9.1) | 0 (0.0) | 1 (10.0) | | |
| Baseline ECOG performance status | | | | | | |
| 0 | 7 (53.8) | 4 (36.4) | 4 (40.0) | 5 (50.0) | N/A | 0.86 |
| 1 | 5 (38.5) | 5 (45.5) | 4 (40.0) | 5 (50.0) | | |
| 2 | 1 (7.7) | 2 (18.2) | 2 (20.0) | 0 (0.0) | | |
| irAE types[b] | | | | | | |
| Arthritis | N/A | 2 (18.2) | N/A | 0 (0.0) | N/A | **0.010** |
| Colitis | | 0 (0.0) | | 3 (30.0) | | |
| Dermatitis | | 0 (0.0) | | 2 (20.0) | | |
| Duodenitis | | 1 (9.1) | | 1 (10.0) | | |
| Gastritis | | 0 (0.0) | | 1 (10.0) | | |
| Hepatitis | | 1 (9.1) | | 4 (40.) | | |
| Hypophysitis | | 0 (0.0) | | 2 (20.0) | | |
| Meningitis | | 0 (0.0) | | 3 (30.0) | | |
| Myocarditis | | 1 (9.1) | | 0 (0.0) | | |
| Myositis | | 2 (18.2) | | 0 (0.0) | | |
| Nephritis | | 1 (9.1) | | 1 (10.0) | | |
| Pancreatitis | | 1 (9.1) | | 1 (10.0) | | |
| Pneumonitis | | 2 (18.2) | | 0 (0.0) | | |
| Thyroiditis | | 1 (9.1) | | 1 (10.0) | | |

*Groups are compared by Kruskal-Wallis (continuous data) or Fisher's exact test (categorical data). Abbreviations: anti-PD-1 denotes 'anti-PD-1 monotherapy', cICI 'combined immune checkpoint inhibition', CTCAE 'Common Terminology Criteria for Adverse Events', irAE 'immune-related adverse event'; N/A 'not applicable', NOTx 'no clinically relevant irAEs', TOX 'with clinically relevant irAEs', yr 'years'. [a] In 2/4 cICI TOX patients and 3/4 anti-PD-1 TOX patients Timepoint 2 sample is missing because irAE(s) developed after one cycle of ICI .[b]Numbers exceed 100% because patients may have more than one irAE simultaneously.*

**Supplementary Table 4. Top 10 parameters contributing to principal component 1 and 2**

| Top 10 unique read-out parameters | Principal component (PC) 1 | Principal component (PC) 2 |
|---|---|---|
| 1 (highest loading score) | granB$^+$ of PD-1$^-$LAG-3$^-$ CD95$^+$ CD8$^+$ T cells | Ki67$^+$ of CXCR3$^+$CCR4$^-$ Th1-associated CD45RO$^+$ memory CD4$^+$ T cells |
| 2 | granB$^+$ of PD-1$^+$LAG-3$^-$ CD95$^+$ CD8$^+$ T cells | Ki67$^+$ of CD4$^+$ T cells |
| 3 | CD45RA$^+$CD27$^+$ naive of CD8$^+$ T cells | Ki67$^+$ of CD45RA$^-$CD27$^-$ effector memory CD4$^+$ T cells |
| 4 | CD57$^+$ of CD8$^+$ T cells | Ki67$^+$ of CD8$^+$ T cells |
| 5 | CD45RA$^+$CD27$^-$ $T_{EMRA}$ of CD8$^+$ T cells | Ki67$^+$ of CXCR3$^+$ CD45RO$^+$ memory CD8$^+$ T |
| 6 | CD4$^+$ T cells of CD3$^+$ T cells | Ki67$^+$ of CD45RA$^+$CD27$^-$ CD8$^+$ $T_{EMRA}$ cells |
| 7 | CD8$^+$ T cells of CD3$^+$ T cells | Ki67$^+$ of CD45RA$^-$CD27$^+$ central memory CD8$^+$ T cells |
| 8 | CD57$^+$ of CD95$^+$ CD8$^+$ T cells | Ki67$^+$ of CD45RA$^-$CD27$^-$ effector memory CD8$^+$ T cells |
| 9 | IFN-$\gamma^+$ of CD4$^+$ T cells | Ki67$^+$ of CD45RA$^-$CD27$^+$ central memory CD4$^+$ T cells |
| 10 (lowest loading score) | IFN-$\gamma^+$ of CD45RO$^+$ memory CD4$^+$ T cells | Ki67$^+$ of CXCR3$^-$CCR4$^+$ Th2-associated CD4$^+$ T cells |

*All parameters represent parental percentages based on manual gating. Each biologically unique population is included only once in the top 10. For example, the percentage CD4$^+$ of CD3$^+$ T cells was assessed in multiple panels and each panel-specific parameter contributed individually to the top 10, but percentage CD4$^+$ of CD3$^+$ T cells is included only once in PC1 top 10. Abbreviations: EMRA denotes 'effector memory re-expressing CD45RA', granB 'granzyme B'.*

### Supplementary Figures

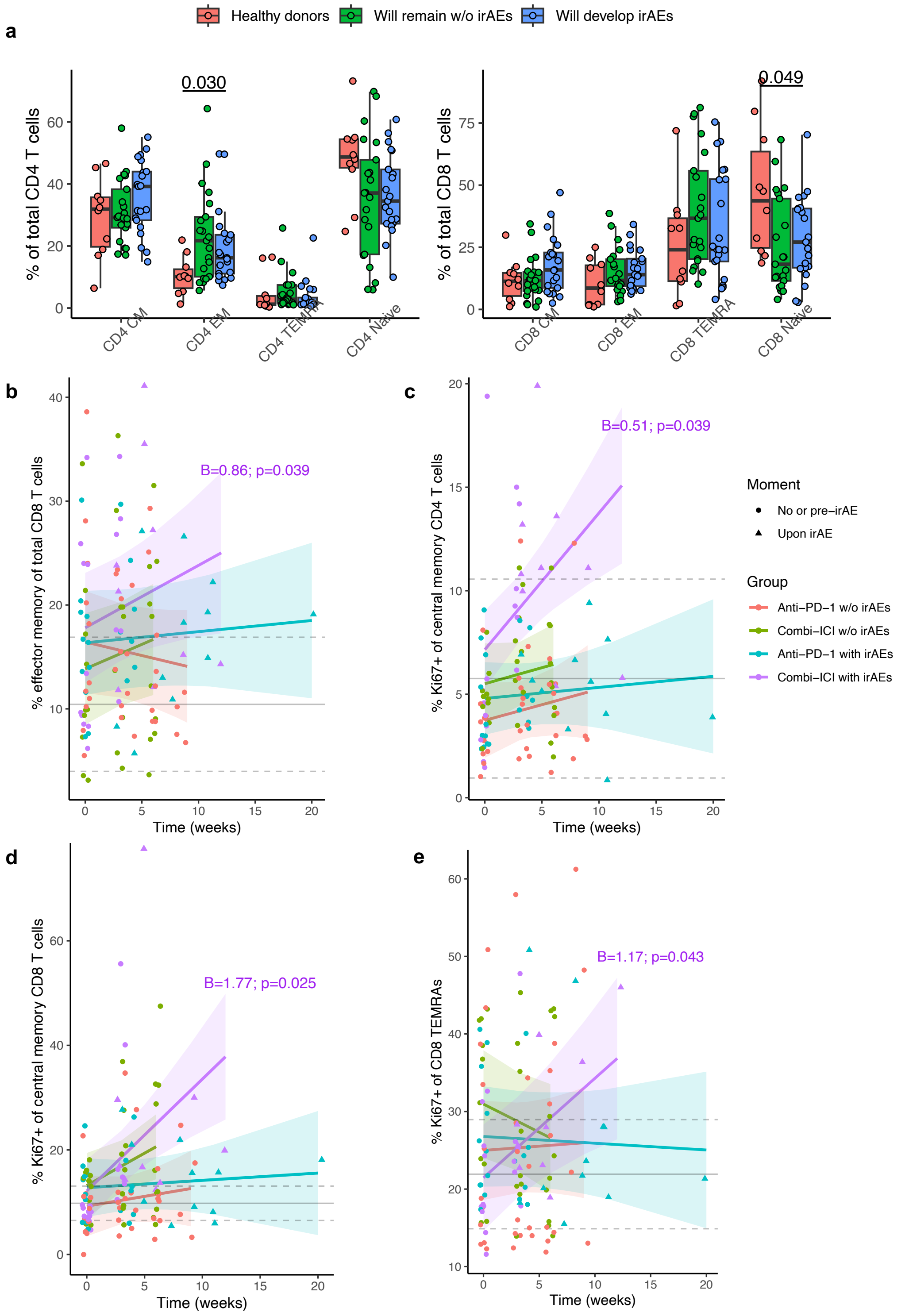


**Supplementary Figure 1 Patients with cancer have different baseline naïve/memory subset distribution than healthy donors, while over time all CD4$^+$ and CD8$^+$ memory subsets in combined-ICI treated patients with toxicity demonstrate enhanced proliferation compared to other patient groups.** (**a**) Baseline abundance of central memory (CM), effector memory (EM), TEMRA and naive subsets in CD4$^+$ T cells (left) and CD8$^+$ T cells (right). Comparisons by Kruskal-Wallis tests. (**b**) Percentage CD8$_{EM}$ of total CD8$^+$ T cells over time. Only significant coefficients for the interaction term with time ('B'), indicating statistically significant change compared to other groups, from mixed-effects models are shown. Gray solid and dashed lines indicate mean healthy donor level with 95% confidence interval. (**c**-**e**) Percentage proliferating of total CD4$_{CM}$ (**c**), CD8$_{CM}$ (**d**) and CD8$_{EMRA}$ (**e**) T cells over time. irAE: immune-related adverse event, w/o: without.

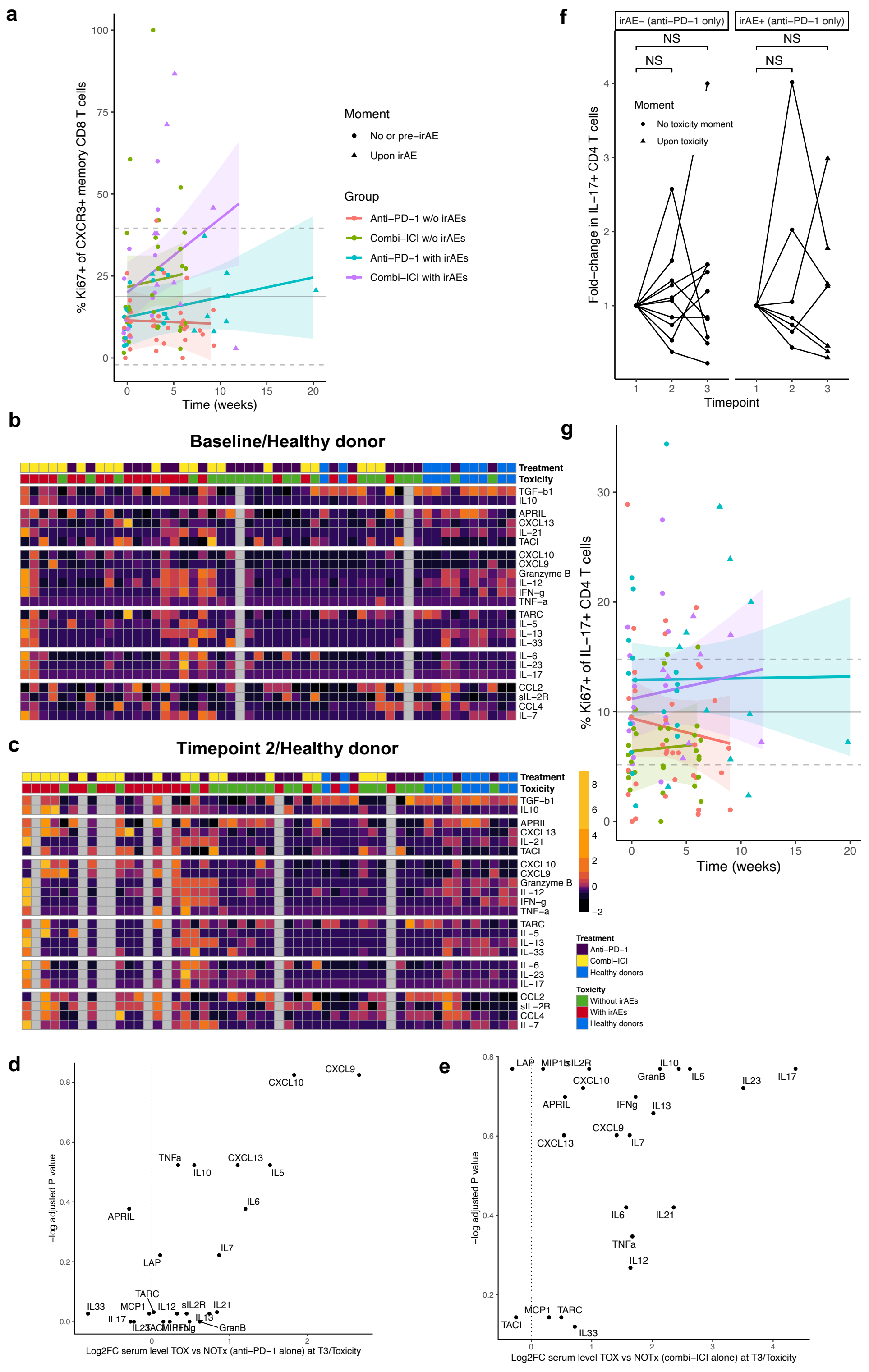

**Supplementary Figure 2 Especially Th1- and Th17-associated cytokines increase towards toxicity after combined ICI, while for anti-PD-1 associated toxicity this is limited to increase in Th1-associated cytokines without changes in IL-17$^{+}$ producing cells compared to patients without toxicity.** (**a**) Percentage proliferating of CXCR3$^{+}$ CD8$^{+}$ T cells over time. Mixed-effects models showed no significant interactions with time. Gray solid and dashed lines indicate mean healthy donor level with 95% confidence interval. (**b**,**c**) Heatmaps showing serum levels of cytokines and chemokines at baseline (**b**) and Timepoint 2 (**c**) or in healthy donors. Data are scaled by individual proteins across timepoints (see **Fig. 3c**), enabling direct comparison of individual proteins between three timepoints. Grey columns indicate missing timepoints; column order (patients) is as in **Fig. 3c**.

(**d,e**) Volcano plots showing relative increase in serum protein levels at Timepoint 3/Toxicity in patients with irAEs (TOX) versus patients without irAEs (NOTx) for (**d**) anti-PD-1 monotherapy and (**e**) combined ICI treated patients. Full protein names are in **Supplementary Table 2**. (**f**) Fold-change in percentage IL-17$^{+}$ of CD4$^{+}$ T cells relative to baseline in anti-PD-1-treated patients without irAEs (left; NOTx) and with (right; TOX) irAEs. (**g**) Percentage proliferating of IL-17$^{+}$ CD4$^{+}$ T cells over time. The same legend, graphical approach and statistics as in (**a**) apply; no significant interactions with time. irAE: immune-related adverse event, NS: not significant, w/o: without.

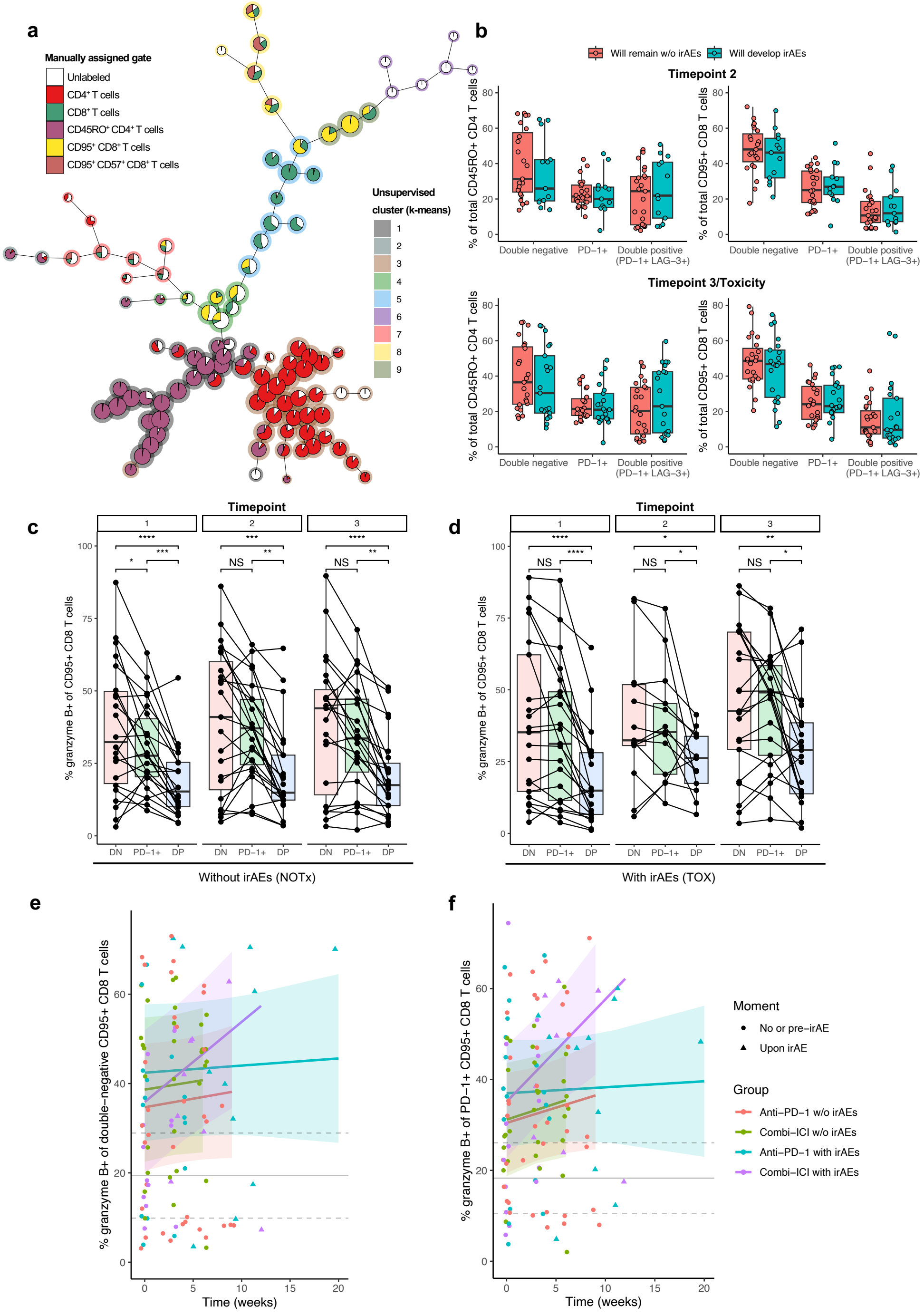


**Supplementary Figure 3 Relative abundance of PD-1$^+$LAG-3$^+$ double positive (DP) CD8$^+$ memory T cells while on-treatment remains unchanged, but towards toxicity especially these DP CD8$^+$ memory T cells show increased cytotoxic potential.** (**a**) FlowSOM visualization created with concatenated file containing 100% of CD3$^+$ T cells from all patients at Timepoint 3/Toxicity and healthy donors, overlaid with k-means clustering results and manually assigned gates. (**b**) Percentages of CD45RO$^+$ CD4$^+$ (left) and CD95$^+$ CD8$^+$ (right) T cells positive for PD-1 alone or PD-1 and LAG-3 at Timepoint 2 (top) or Timepoint 3/Toxicity (bottom). (**c**,**d**) Percentages of CD95$^+$CD8$^+$ T cells producing granzyme B over time after PMA/ionomycin stimulation, depending on inhibitory receptor expression pattern, in patients without (**c**) or with irAEs (**d**); anti-PD-1- and cICI-treated patients are combined. Comparisons by pairwise paired Wilcoxon tests with Benjamini-Hochberg correction for multiple testing. (**e**,**f**) Percentage of granzyme B producing (**e**) PD-1$^-$LAG-3$^-$ double-negative and (**f**) PD-1$^+$ CD95$^+$CD8$^+$ T cells over time analyzed by mixed models; no significant interactions with time. Gray solid and dashed lines indicate mean healthy donor level with 95% confidence interval. irAE: immune-related adverse event, NS: not significant, w/o: without. * $P<0.05$, ** $P<0.01$, *** $P<0.001$, **** $P<0.0001$.

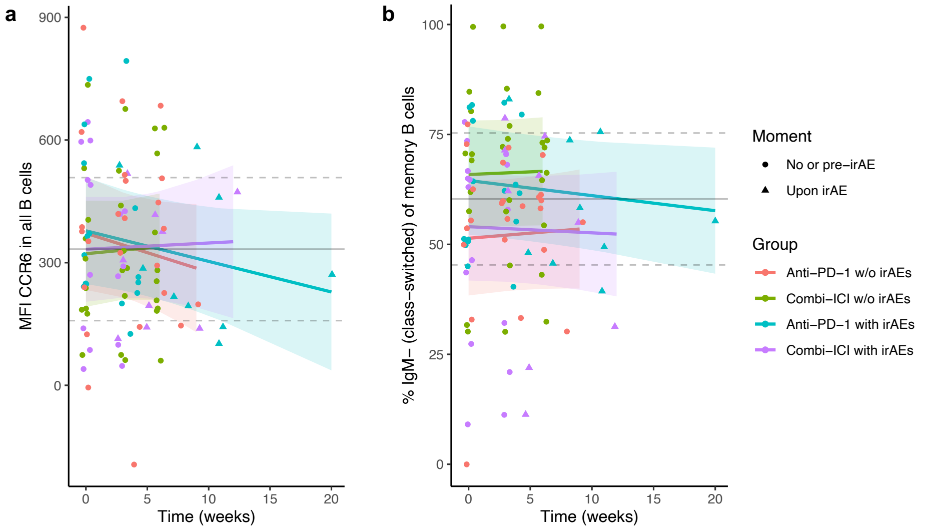


**Supplementary Figure 4 Extent of B cell activation and class-switching do not change among groups over time.** (**a**) Median fluorescence intensity (MFI) of CCR6 in all B cells over time and (**b**) Percentage IgM$^-$ class-switched of CD27$^+$ memory B cells over time. Mixed-effects models showed no significant interactions with time. Gray solid and dashed lines indicate mean healthy donor level with 95% confidence interval. irAE: immune-related adverse event, w/o: without.

**a**

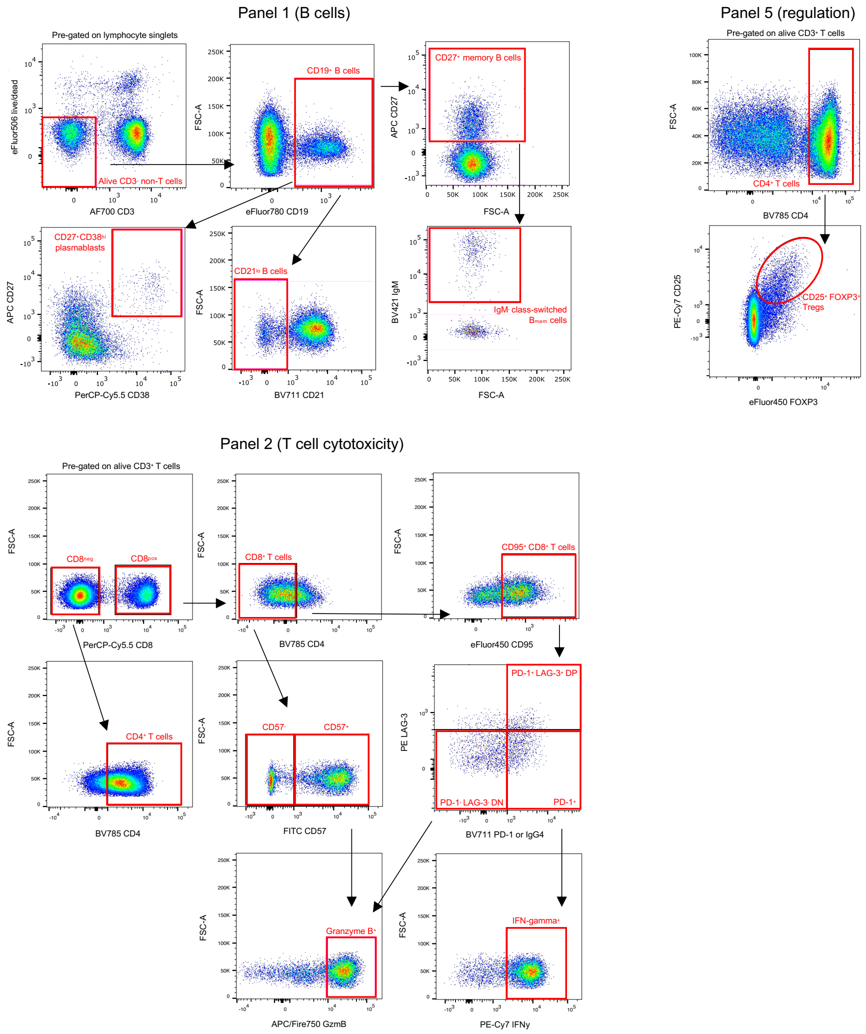

**b**

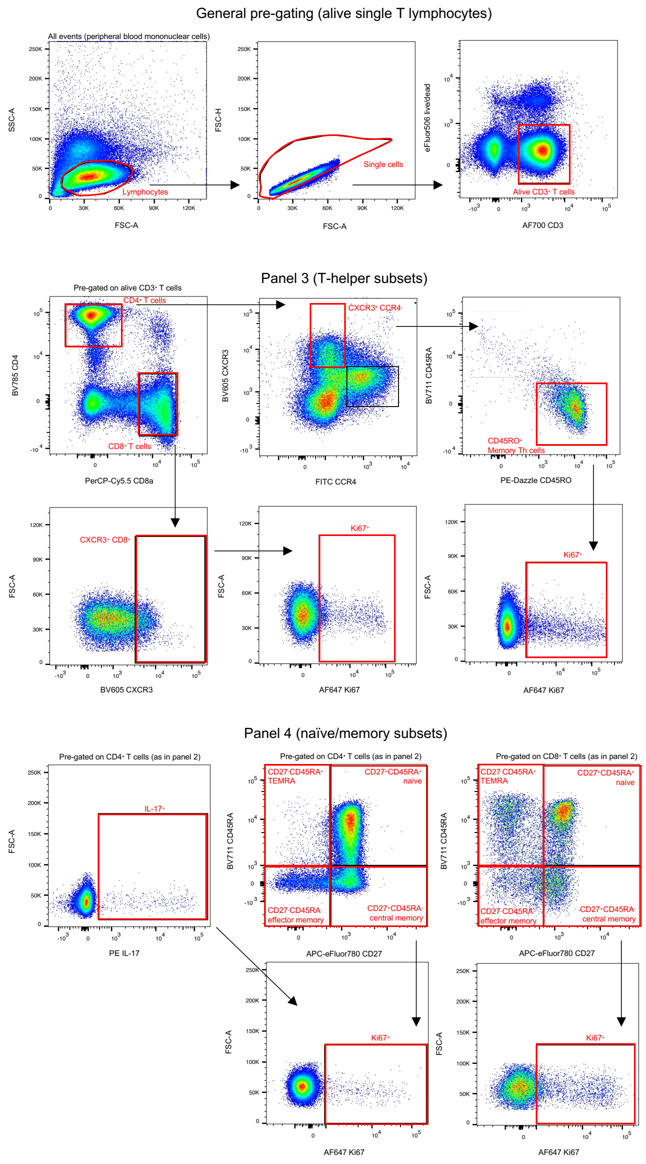


**Supplementary Figure 5 Representative flow cytometry plots illustrating gating strategies.** (**a**) Panel 1 (B cells), Panel 2 (T cell cytotoxicity) and Panel 5 (regulation). Pre-gating strategy for Panels 2 and 5 is in (**b**). (**b**) General pre-gating (alive single T lymphocytes), Panel 3 (T-helper subsets), Panel 4 (naïve/memory subsets).
